# Perceived barriers and facilitators to implementing preventive therapy for tuberculosis in Malaysian prisons: Perspectives from prison personnel

**DOI:** 10.1101/2024.11.05.24316771

**Authors:** Phoebe Chen, Frederick Altice, Divya Ghoshal, Ahsan Ahmad, Jiang Long, Rumana Saifi, Hui Moon Koh, Naga Nagadevi, Mohd Norbayusri Baharudin, Esther Wong Min Fui, Adeeba Kamarulzaman, Sheela Shenoi

## Abstract

Tuberculosis (TB) is a major health threat worldwide. TB is concentrated in prisons, where TB preventive therapy (TPT) implementation has been limited. Prisons generally, and especially in Malaysia, are overcrowded, minimally resourced, and have high prevalence of latent TB.

As prison personnel are largely responsible for TPT implementation, their perspectives were sought to identify barriers and potential solutions for implementing TPT in Malaysia’s largest prison. A focus group using nominal group technique (NGT) was conducted with 9 prison officers with experience related to TPT to generate a list and rank-order the top TPT barriers. The process was repeated to assess facilitators.

The top three barriers to implementing TPT were: 1) disruptions involved in prison activities (e.g., intra- and extra-prison transfers; 8 votes); 2) insufficient workforce (5 votes), and unanticipated inter-prison transfers (4 votes). Potential solutions included implementing care coordination protocols between officers and medical staff (8 votes), placing a medical hold until TPT completion (5 votes), and improving medication delivery logistics (4 votes). All prison officers supported a short-course TPT regimen administered once-weekly for 12 weeks (i.e. rifapentine and isoniazid (3HP)), over a daily TPT regimen for 6 months (i.e. isoniazid (26H)), due to less logistics and staff time required.

From the perspective of prison officer stakeholders, logistical constraints commonly observed in prisons were perceived to impact the implementation of TPT, yet they prioritized pragmatic solutions that align with known implementation strategies to employ to optimize TPT delivery, including developing stakeholder relationships between medical and prison personnel and changing infrastructure to address frequent transfers within and between prisons and the community.

## Introduction

Tuberculosis (TB) is the world’s second deadliest infectious disease, second only to COVID-19, despite the existence of highly effective treatment and preventive therapies [1]. A quarter of the global population is estimated to have latent TB infection (LTBI) and are at risk of developing active TB disease [2]. TB disproportionately impacts incarcerated populations, who have a disease burden ten times higher than the general population [3], due to factors like overcrowding, malnutrition, and delayed diagnosis [4]. In Malaysia, over a quarter of the population has LTBI [5], with significantly higher prevalence in Malaysian prisons (68% to 88%), as identified previously [6–8]. TB prevalence is concentrated in prisons and complicated by overcrowding, punitive drug policies, and limited prison resources [4,9]. Structural challenges to TB control in prisons include vulnerable and marginalized populations pre-disposed to high TB risk, inadequate ventilation, poor nutrition, comorbid substance use, and poor access to medical care [10]. Periods of incarceration thus present an opportunity to intervene in the TB epidemic by targeting vulnerable individuals during a critical time period of heightened risk [11]. Even though controlling communicable diseases, including TB, among incarcerated persons is included in Malaysian prison regulations [12], prison management faces difficulties in controlling the disease in the high risk prison environment, hence impacting institutions and creating pressures on treatment quality, incident risk, and health of both staff and incarcerated persons [13].

TB preventive therapy (TPT) is a safe, highly efficacious, and cost-effective method to prevent the progression of latent TB infection (LTBI) to active TB disease [2]. TPT is one of the key pillars of WHO’s End TB Strategy to control the worldwide TB epidemic [2]. In 2018, at the UN High-Level Meeting on TB, all country leaders pledged to provide TPT to at least 30 million people at risk of progression to TB disease by 2022; however, only 15.5 million people received TPT between 2018 and 2022 [3]. Despite advances in shorter and better tolerated TPT regimens and reduced costs, TPT remains underutilized globally [14]. The WHO considers the scale-up of TPT to be critical to the End TB Strategy, and UN leaders recently pledged to provide TPT to 45 million people by 2027 [15]. Challenges to scaling up TPT include poor program performance in identifying opportunities to test and treat target populations, limited uptake of shorter TPT regimens, and cost [15,16]. TPT remains unavailable in most prisons, despite a strong case for implementation in this high risk and high prevalence setting. As of 2023, the TB burden in prisons is substantial and growing globally [14]. Prisons serve as important reservoirs for TB, including multi-drug-resistant (MDR) TB, which is transmitted to the general population through the prison workforce and release of incarcerated persons [11,17–19].

To scale up TPT in prisons, a better understanding of the implementation context is required. To date, there is limited research informing TPT implementation in low- and middle-income countries (LMICs), particularly within prisons [17,20–24]. It is difficult to implement TPT in the prison environment without relevant stakeholder input on context and facilitators. Therefore, it is crucial to obtain insights from prison staff who would administer and coordinate TPT initiatives. This study elicited prison officers’ perspectives on TPT barriers and facilitators in Malaysia’s largest prison, which was vital to filling in the evidence gap using an established implementation framework.

## Methods

To understand the barriers to implementing TPT in prisons in Malaysia, we used the established Practical Robust Implementation and Sustainability Model (PRISM), which expands the Reach, Effectiveness, Adoption, Implementation, Maintenance (RE-AIM) framework to identify contextual factors to guide our pre-implementation process. Since multiple factors may influence TPT implementation, we examined the inner context by interviewing prison officers who are key implementers and can either impede or promote TPT adoption and implementation. In Malaysia, the prevalence of latent TB among prison officers is approximately 81%, as high as it is for people in prison, making prisons a high-risk environment to anyone in this setting [25]. As such, they are key stakeholders for whether and how TPT is implemented.

### Participants and Setting

In July 2023, we recruited 10 participants from Malaysia’s largest prison near Kuala Lumpur using purposive sampling. The prison director introduced the study and invited 10 prison officers to participate. Eligibility included employment at the prison for at least 6 months and had direct experience interfacing with medical staff and assisting with medication administration. Prison officers had no formal TB or TPT training, though they possessed a basic understanding of TB informally passed on from prison medical staff. The study was voluntary and conducted in a private room outside the prison with one facilitator and one translator for English and Bahasa Malaysia. One participant generated ideas but had to leave before voting, leaving 9 participants who contributed data to the rank-ordering.

Kajang Prison houses over 5000 incarcerated persons [26] and is overcrowded, making incarcerated persons vulnerable to TB infection. The medical unit employes 4 physicians and 6 medical assistants, with a doctor to patient ratio of 1:1250. All persons newly-admitted to prison undergo screening for HIV;while national guidelines also recommend screening for TB, implementation lags [27]. If a person has symptoms observed by a prison officer or healthcare provider, they are isolated, evaluated, and, if testing positive for active TB, initiated on treatment at a nearby government hospital, with the remainder of treatment to be completed in the prison. There is no standardized protocol for medication administration across prisons. In Kajang prison, the prison officer collects medication from the medical clinic and is responsible for administering daily observed therapy in the TB housing unit. TPT is not currently deployed.

### Procedures

Participants completed a brief survey prior to the session and provided verbal consent and were reminded that the discussion was confidential. No compensation was provided. The study protocol was approved by the Institutional Review Boards at Yale University (#2000020053) and the Universiti Malaya (#17-19-33974), and the Medical Research & Ethics Committee (MREC) at the Malaysian Ministry of Health (#NMRR ID-17-19-33974), with approval from the Malaysian Prison’s Department.

The focus group was conducted using nominal group technique (NGT). The expert facilitator interacted with participants while research assistants took notes and recorded responses on a whiteboard for all participants to observe. The session was audio-recorded, transcribed, and translated.

NGT is a well-established mixed-methods strategy increasingly used in implementation research to rapidly identify barriers and solutions for health service delivery [27–31]. We chose NGT to obtain both quantitative estimates of priority (rank-ordering) and in-depth qualitative information that could be rapidly implemented. Compared to traditional focus groups, NGT also ensures more equitable participation from all participants. The NGT process was as follows: (a) silent generation of ideas in response to the initial question “What gets in the way of prisoners receiving medications to prevent TB every day for 6 months?”; (b) round-robin feedback from participants where each participant contributed a single idea on a whiteboard until saturation was reached; (c) clarification of ideas as needed by the moderator, allowing for structured group discussion and evaluation of ideas; some ideas were grouped together by consensus and duplicate items were removed; and (d) individual voting, with each participant allocating up to 3 votes to idea(s) they deemed most important. Votes were tallied and the group decision was mathematically derived through rank-ordering [27]. The facilitator led a final discussion to review the participants’ results and ensure that they had face validity among participants. This process was repeated with the second question “What could you do or what would be needed to fix these problems that get in the way?” to elicit proposed solutions based on the top-3 ranked barriers. Last, the facilitator proposed a new idea to “Change to a weekly (not daily medication) for TPT” and asked participants with this new information, how many of their 3 votes, if any, they would re-allocate to this idea.

Focus group questions were based on prior literature and experts in TB treatment, NGT methodology, and prison health.

## Results

### Demographics

All participants (n=10) were male prison officers whose mean age was 32.7 years and had worked as a prison officer for a mean of 8.6 years. Nine respondents (90%) voted on both questions. 1 participant contributed to the discussion but left prior to the first round of voting due to urgent prison duties.

### Perceived Barriers

**Table 1** presents perceived barriers to implementing TPT in the Malaysian prison setting. The top three barriers to TPT delivery were interruptions to the routine prison schedule (8 votes), including transfers within the prison and to outside appointments (e.g. due to court cases, hospital visits, transfers within prison housing units), limited prison staff to ensure TPT delivery (5 votes), and unanticipated transfer to another prison (4 votes). The remaining barriers are listed in Table 1.

**Table 1:**
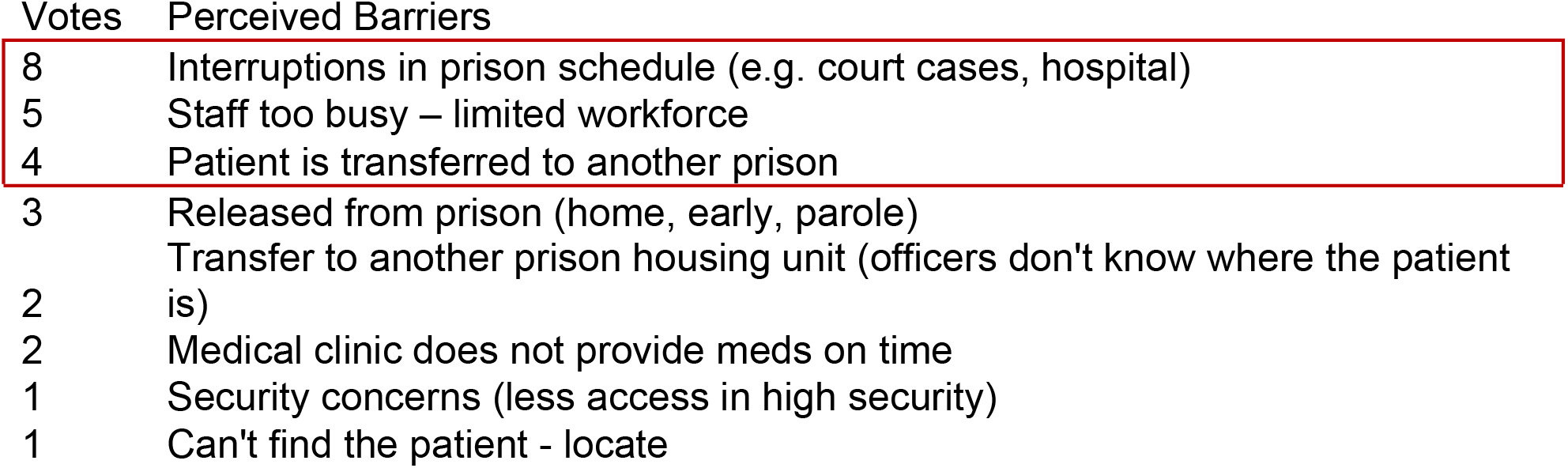
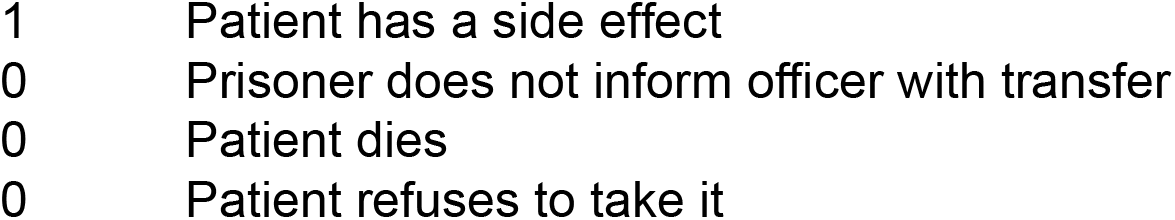
Key barriers for implementing TPT in prison (N=9)

### Perceived Facilitators

Table 2 presents perceived facilitators for the 3 main barriers identified. The top three solutions were improving care coordination between prison officers and medical staff to ensure continued medication delivery when schedule disruptions occurred (8 votes), deploying a medical hold to avoid transferring the patient until TPT was competed (5 votes), and moving medications closer to patient housing units with a supply of medication at each prison housing unit (4 votes). The remaining facilitators are listed in Table 2.

**Table 2:**
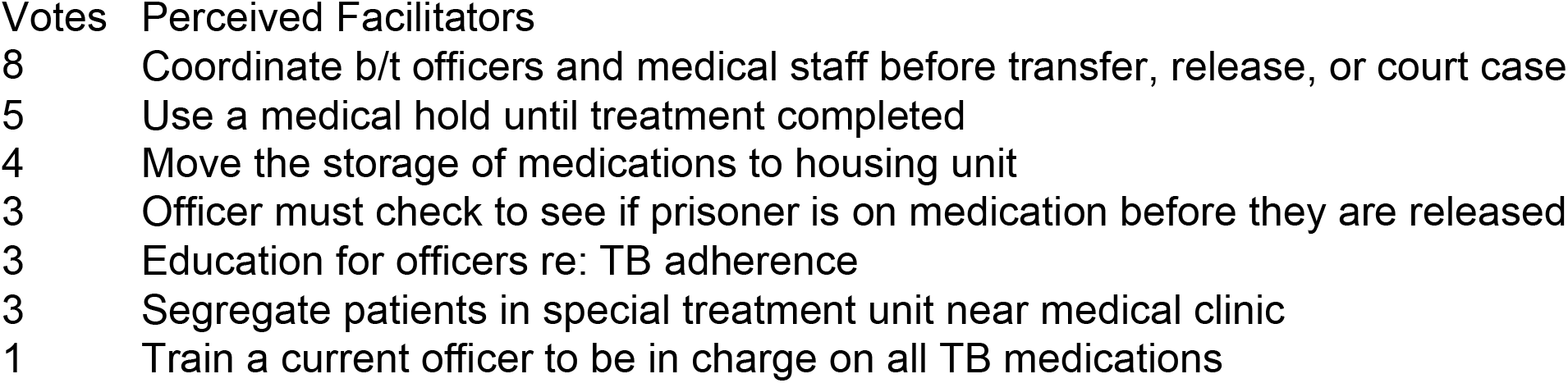
Key perceived facilitators for initiating TPT in prison.

| Votes | Perceived Facilitators |
| --- | --- |
| 8 | Coordinate b/t officers and medical staff before transfer, release, or court case |
| 5 | Use a medical hold until treatment completed |
| 4 | Move the storage of medications to housing unit |
| 3 | Officer must check to see if prisoner is on medication before they are released |
| 3 | Education for officers re: TB adherence |
| 3 | Segregate patients in special treatment unit near medical clinic |
| 1 | Train a current officer to be in charge on all TB medications |

Finally, when presented with an option at the end of the NGT session to “Change to a weekly (not daily medication)”, participants re-allocated 19 votes of 27 votes to this option. The majority of participants expressed this change would dramatically improve adherence and maximize use of their limited human resources.

## Discussion

As Malaysia struggles to control its TB epidemic in a country with high TB burden, it is crucial to target TPT to those most vulnerable and accessible – to people in prison where TB prevalence is extraordinarily high (68% to 88%) [6–8]. There have been recent calls for accelerated strategies to screen for TB [10] and to introduce TPT in prisons [14], yet guidelines on how to best deliver TPT in this setting require tailoring to each context [32].

In the absence of clear guidelines for TPT delivery in prisons, the PRISM/RE-AIM framework provides an ideal heuristic for pre-implementation assessment to guide future implementation. Though people in prison are the major target for TPT, their access to treatment can be hindered or facilitated by multiple stakeholders, including prison medical staff and custodial staff. We used this framework to prepare for a future TPT implementation program in prisons, and, in collaboration with prison officer stakeholders, explore potential implementation strategies based on themes prioritized by prison personnel [33–37].

Though many of the barriers and facilitators to TPT may be common in prisons, it is important to tailor implementation to context. For example, in Malaysia, prison turnover has substantially increased with shortened stays in prison and multiple disruptions with transfers between housing units, prisons and to the community either for court appearances or medical appointments at government hospitals or release to home.

Important here is that the prison officers were unaware of newer TPT options like 3 months of once-weekly isoniazid plus rifapentine (3HP). They perceived that switching to this strategy rather than the standard 6 months of daily isoniazid preventive therapy (IPT) would minimize disruptions in medication delivery due to typical and sometimes unexpected logistical transfers. This short-course regimen with less frequent dosing has higher acceptability and completion rates relative to IPT and may be especially useful in prisons where high turnover logistical barriers undermine treatment completion (14,17,20,22,37). Recent TPT programs in prisons in Thailand and Pakistan achieved 87-93% completion rates [20,21]. A prior systematic review of 6-month IPT completion rates in prisons was substantially lower at 44%, reducing the enthusiasm for such a strategy by international agencies [17]. Cited barriers by prison personnel was the shortage of staff, insufficient knowledge and training, and a lack of prioritization for prison health and resources by governmental authorities [38,39], which might partially be solved by a once-weekly, 3HP short-course therapy that would reduce work demands on staff and minimize security demands by minimizing movement by people in prison. In addition, re-entry to the community and prison transfers are known high-risk events for interruption in TB care.

### Develop stakeholder inter-relationships

In the absence of a 3HP strategy, prison officers’ top suggestion to optimize TPT was to enhance care coordination between prison officers and medical staff, which could be accomplished through local consensus discussions and checklists [10,23,40]. For example, prison officers could meet with medical personnel at the end of each week to discuss any anticipated transfer expected for the following week. In such cases, medical staff might try a number of approaches like prepare transfer medications to people who are moving to another prison or deliver medications the day before for offsite visits. As there can be a tension between medical and custodial staff in terms of prioritizing health vs security, creating a shared vision for creating a safe environment to minimize TB transmission to personnel might align their missions. This is especially true in places like Malaysia where in the absence of such a shared mission, prison personnel have latent TB infection rates similar to people in prison due to a shared risk environment [25].

### Change infrastructure

The second-ranked implementation strategy supported by prison officers was to change prison infrastructure to minimize disruptions due to transfers. Key among them, which has been done in other prison settings, is to ensure that the TPT medications are closer to housing units. instead of at the medical clinic, and provision of a temporary medication supply accompanying the patient during transfers or release to finish their treatment course. Though they did not provide specifics, this infrastructure change might include the entire week’s supply of medication being delivered to the housing unit for dispensation, or in the case of daily observed therapy, having the medical staff make daily deliveries to the housing unit rather than rely on a traditional “roll call” where people in prison must leave their units to receive medications at the medical unit. An alternative, but equally important suggestion was to have the custodial staff institute a “medical hold” for any patients prescribed TPT, which would prevent such persons from any kind of transfer until there was secondary clearance by a medical officer.

### Training and Education

Though identified as a lower priority, prison officers also identified the need for more trainings and knowledge about TB and TPT. Such a strategy could have benefits that include creating a shared vision of reducing the risk for TB transmission to prison personnel, but also to reduce TB incidence nationally in Malaysia. In many prisons, including in Malaysia, only prison medical staff receive TB training from the Ministry of Health. These trainings emphasize active TB and do not adequately address latent TB or TPT. Unilateral training undermines any shared alignment for ensuring TPT. Formal comprehensive training and knowledge-sharing on TB and TPT between medical and non-medical prison staff could bridge the knowledge gap and emphasize the importance of TPT and medication adherence to all personnel. In low-resource settings, educational meetings and training workshops have strong evidence for improving patient outcomes. For example, in Thai prisons, high health literacy among prison officers was associated with good TB practices [42,43]. The U.S. Centers for Disease Control and Prevention (CDC) also recommends all correctional officers in high risk facilities receive TB training [43]. Further data are needed on knowledge level of TB and TPT and training needs for prison staff but should be explored further.

Of interest here is the lack of discussion or prioritization of other key stakeholder relationships that might influence TPT delivery, like with the Ministry of Health and prison leadership. Political will is crucial for a successful response to TB in prisons [11,24,40]. This political will not only requires executive sponsorship from leadership, but also a financial commitment to prioritize TPT implementation in prisons [40]. In the case of prisons, it often requires a joint commitment by both the Ministry of Justice (or sometimes Interior) and Ministry of Health and unless both are fully committed to prioritize TB control, then implementation efforts might not be fully optimized. Stakeholder engagement and consensus-building will be important to bridge conflicting priorities for criminal justice (security) and public health (prevention and treatment) [11,40]. Such aligned engagement would be crucial to optimizing the national budget for TB and ensure that a proportionate amount is allocated to TB control in prisons. Given the limited amount of funding allocated to prisons and health, blended implementation strategies that include a combination of low threshold strategies like education and infrastructure changes as suggested by the officers may be low cost, though care coordination and evaluative and iterative strategies may be needed to change day-to-day operations. Such examples can include audit and feedback to ensure that all people who enter prisons are screened for latent TB infection, and if diagnosed, that there are methods to track TPT initiation and adherence to treatment.

This might involve creation of checklists to diagnose latent TB infection or dashboards that track the number of people on treatment and notifying prison officers of adherence rates with real-time feedback cycles.

### Study Limitations

While there are many important and pragmatic perspectives listed and ranked by prison officers, this sample represents only one stakeholder group - the prison officers who control the movement of people in prison. As such, they represent one important gatekeeper to implementing TPT. Their perspective should be reviewed and triangulated with other stakeholders as was done recently when identifying and prioritizing barriers to implementing syringe services programs in prison in Canada [29,44]. Future studies also need to involve other stakeholders, like clinicians in prison people in prison, and outer context authorities.

While a single NGT session might be insufficient to be fully representative or achieve saturation, the findings here have face validity and accurately represent known barriers. Their solutions, however, might represent an initial plan to select and prioritize implementation strategies and follow-up NGT could be done to determine if the observed barriers were successfully overcome.

Lastly, while the barriers and facilitators represent the perception by the officers, they may not be fully accurate. For example, in this session, a substantial number of officers changed their vote after learning about an innovative TPT strategy – a 3-month TPT regimen that requires administration only once per week. Thus, in the absence of being informed by ideas outside the prison, their selection of implementation strategies may not fully align with all the true possibilities. Notwithstanding these limitations, the ideas generated provide a heuristic for implementing actionable changes to optimize TPT delivery in Malaysian prisons.

## Conclusions

Despite several decades of TPT and strong evidence supporting its efficacy, implementation of TPT in prisons has been limited with challenges in adherence and completion rates. However, prisons are sentinel sites for TB reactivation, transmission, and development of MDR TB with dire public health consequences. Prioritizing more stakeholder engagement, changes to infrastructure and logistical processes, and resources to optimize TPT implementation in prisons may greatly improve TB control. In the future, this pre-implementation framework may be a useful model for other prisons in Malaysia and other low-resource settings.

## Data Availability

The data are included in the manuscript.

## Acknowledgements

We appreciate support for this project from the Malaysian Ministry of Health, Prisons Department, and Centre of Excellence for Research in AIDS, University Malaya.

